# Utility of polygenic risk scores in diagnostic genetic testing for dilated cardiomyopathy

**DOI:** 10.64898/2026.01.20.26344484

**Authors:** Utpala Nanda Chowdhury, Eleni Giannoulatou, Diane Fatkin, John J. Atherton, Nathan J Palpant, Baptiste Couvy-Duchesne, Sonia Shah

**Affiliations:** Institute for Molecular Bioscience, The University of Queensland, St Lucia, QLD, Australia; Department of Computer Science and Engineering, University of Rajshahi, Rajshahi, Bangladesh; Victor Chang Cardiac Research Institute, 405 Liverpool St, Darlinghurst, New South Wales 2010, Australia; School of Clinical Medicine, Faculty of Medicine and Health, UNSW Sydney, Kensington, New South Wales, Australia; Cardiology Department, St Vincent’s Hospital, Darlinghurst, New South Wales, Australia; Faculty of Health, Medicine and Behavioural Sciences, The University of Queensland, St Lucia, QLD, Australia; Cardiology Department, Royal Brisbane and Women’s Hospital, Herston, QLD, Australia; Brain & Mental Health Program, QIMR Berghofer Medical Research Institute, Brisbane, QLD, Australia; School of Biomedical Sciences, Queensland University of Technology, Brisbane, Australia

**Keywords:** Dilated cardiomyopathy, Genetic testing, Polygenic risk scores, *TTN* truncating variants

## Abstract

Dilated cardiomyopathy (DCM) is a major driver of heart failure and transplantation. Despite its strong genetic basis, clinical genetic testing identifies pathogenic/likely pathogenic (P/LP) variants in only about one-third of cases. Genome-wide association studies (GWAS) indicate that common variants also contribute to genetic risk, but utility of polygenic risk score (PRS) for diagnostic testing is unclear. We derived a DCM PRS from a large GWAS and applied it to 113 Australian Genomics Health Alliance (AGHA) participants of genetically determined European ancestry with clinically diagnosed DCM, with most having a family history of disease. Genetic testing identified a P/LP variant in 34% of patients. Using 450,280 UK Biobank participants of European ancestry as a reference, we selected the best performing PRS (SBayesRC; odds ratio per SD increase in PRS = 1.72, 95% CI 1.58–1.88). We compared the PRS of individuals with a P/LP variant in the *TTN* gene (*TTN_*G+) or other genes (non*TTN_*G+), and in those without any P/LP or variants of unknown significance identified (G-). Relative to the UK Biobank reference, median PRS values were significantly higher in G- and *TTN*_G+ groups. Twenty-six percent of G- and 25% of *TTN*_G+ cases were in the top decile, versus 11% of non*TTN*_G+ cases. Notably, *TTN*_G+ cases were absent from the lowest four deciles. These findings support a substantial polygenic contribution in DCM patients without identifiable monogenic variants as well as in *TTN*-related DCM. Incorporating PRS alongside routine genetic testing could increase diagnostic yield and inform risk management for patients and families.

## 1. Introduction

With a prevalence of approximately 1 in 250 individuals, dilated cardiomyopathy (DCM) is a major cause of heart failure requiring cardiac transplantation [1, 2]. Until recently, familial DCM was considered to be exclusively monogenic, whereby a single large effect rare variant in a single gene is the primary driver of disease susceptibility. Rare pathogenic variants for DCM have been identified in genes encoding desmosome proteins (*DSP, PKP2*), ion channels (*SCN5A*) and sarcomere proteins (*TTN, MYH7, MYH6, MYPN, TNNT2 and TPM1*) [3]. However, these often display incomplete penetrance and expressive variability [4, 5]. Titin truncating variants (*TTNtv)* are the most frequently observed variants, accounting for around 25% of familial DCM cases [6], but are also observed in ∼1.0% of the general population without DCM [7, 8]. Variability in penetrance and expressivity of pathogenic variants is thought to be attributed to environmental stressors and additional genetic factors [9–11].

Genetic testing in patients with a clinical diagnosis of DCM is useful for confirming diagnosis, understanding disease prognosis, informing clinical management, and understanding risk in first-degree relatives. For example, *LMNA* variants are associated with an adverse prognosis and may require more frequent surveillance and earlier consideration of implantable cardioverter defibrillators [12]. The current approach to diagnostic genetic testing involves targeted sequencing of a panel of known DCM genes to identify pathogenic/likely pathogenic (P/LP) variants. However, the diagnostic yield for DCM (i.e. proportion of individuals where a P/LP variant can be identified) is reported to be around 15-25% amongst unselected patients with DCM, and 20-40% in patients with familial DCM [3]. Therefore, for the vast majority of patients with DCM, the underlying cause of disease remains unknown.

Recent genome-wide association studies (GWAS) have revealed a substantial contribution of common, small-effect variants to diseases such as familial hypercholesterolemia (FH) [13], DCM [14, 15] and hypertrophic cardiomyopathy (HCM) [16], supporting a polygenic contribution. A polygenic risk score (PRS) quantifies the cumulative effect of these numerous common risk variants, providing an estimate of an individual’s genetic liability to disease. Previous studies have shown that a high PRS can confer the same disease risk as monogenic disease variants [17, 18]. For example, monogenic FH variant carriers have on average LDL-C levels that are ∼30mg/L higher than individuals that do not carry any FH variants, while individuals in the top 5% of the PRS distribution for LDL-C also have on average ∼30mg/L higher LDL-C levels compared to the rest of the population [19]. Monogenic variants for FH also confer an up to 3-fold risk of coronary artery disease (CAD), which is similar to risk of CAD in individuals with a PRS in the top 8% of the population distribution of the CAD PRS [17], while individuals in the top 10% of the QT-interval PRS distribution have ∼4-fold risk of genotype negative Long-QT syndrome [20]. Several studies have demonstrated PRS contribution in DCM. A GWAS of DCM showed that individuals in the top percentile of the PRS have 4-fold increased risk than the median and 7-fold increased risk compared to the bottom percentile [14]. Another DCM GWAS reported significantly lower mean PRS in P/LP carriers compared to non-carriers and stronger enrichment of P/LP non-carriers amongst those with a high PRS [15]. A study of familial DCM found PRS to be significantly higher in both P/LP carriers and non-carriers compared to a healthy elderly cohort [21]. In addition, PRS can modify the penetrance of pathogenic variants to affect disease age-of-onset, severity and progression [22–25]. Studies on FH were some of the first to demonstrate this. A high PRS for LDL-cholesterol is reported to be the likely cause of disease in around 30% of patients with a clinical diagnosis of FH in whom a monogenic variant cannot be identified [26]. LDL-C PRS testing, in addition to monogenic variant testing for FH, is already implemented in some lipid clinics in the United Kingdom [27]. Reports indicate that patients who receive a probable genetic cause for disease (either monogenic or polygenic) believe more strongly in the use of cholesterol-lowering medication to control their risk and feel less guilty about their lifestyle being the cause of disease [27, 28]. PRS testing, in addition to identification of monogenic variants, can therefore increase the diagnostic yield of genetic testing and have beneficial psychosocial impact on patients.

Recent European Society of Cardiology (ESC) guidelines for the management of cardiomyopathies recommend offering cascade genetic testing to adult at-risk family members only if a P/LP variant has been identified in an affected individual in the family [12, 29]. However, the absence of an identified P/LP variant does not rule out a genetic contribution, as patients may harbour a novel variant not detected by current clinical genetic testing panels or as demonstrated for FH, a high polygenic burden. There is growing evidence that, among affected individuals with a low disease PRS, the likelihood of carrying a monogenic variant is higher [30]. By analysing exome array data in UK Biobank (UKB), Lu et al. reported that a standard deviation decrease in PRS was associated with an almost 3-fold increased odds of identifying rare pathogenic variants [31]. Thus, for individuals where a P/LP variant has not been identified through genetic testing using a panel of known disease genes, a patient’s PRS could provide a cost-effective strategy to prioritise individuals for more comprehensive testing using whole-genome sequencing.

Despite advances in our understanding of the polygenic contribution to DCM, clinical genetic testing in DCM patients focuses solely on identification of P/LP variants in known DCM genes. In this study, PRS analysis in familial DCM patients from the Australian Genomic Health Alliance (AGHA) Cardiovascular Flagship [32] who had undergone clinical genetic testing, was carried out. Recognising that polygenic risk may act in concert with monogenic variants, this study aimed to determine whether knowledge of PRS in patients with and without an identified P/LP variant could be informative for clinical management.

## 2. Materials and Method

### 2.1. Study design

The overview of the study design has been illustrated in Figure 1.

**Figure 1:**
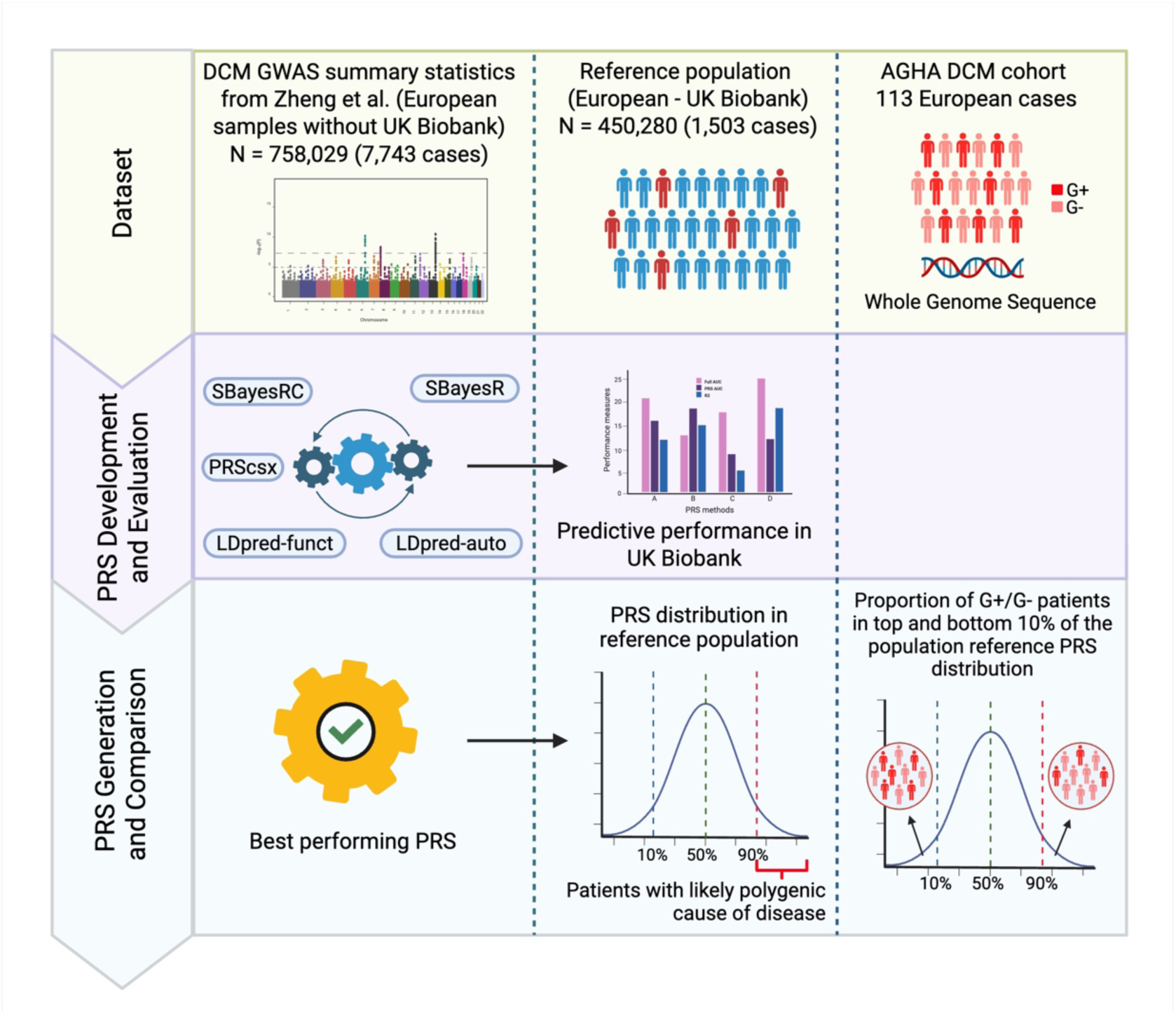
Study overview. The DCM PRS was generated using summary statistics from a European-ancestry DCM GWAS by Zheng et al. [14], not including UKB samples. Performance of five PRS methods for predicting DCM diagnosis was evaluated in European individuals from UKB. The best performing method was used to calculate PRS of the AGHA DCM patients. By overlaying PRS of AGHA DCM individuals of European ancestry onto the UKB European PRS reference distribution, AGHA patients with a high (top decile of the reference distribution) or low PRS (bottom decile) were identified. AGHA - The Australian Genomics Health Alliance cardiovascular flagship cohort. The figure is created with BioRender.com.

### 2.2. Study population – The Australian Genomics Health Alliance cardiovascular flagship (AGHA) cohort

The AGHA Cardiovascular Flagship cohort comprises 600 individuals with a clinical diagnosis of an inherited cardiac disease [32]. Access was provided to whole genome sequence and standardised clinical phenotypic data for 429 individuals (195 female and 234 male) (Table S1), who consented for analysis through Australian Genomics, of which 129 individuals had a clinical diagnosis of DCM (Table 1). DCM was defined by left ventricular end-diastolic diameter more than 112% of predicted and left ventricular ejection fraction less than 45%. DCM cases were included in the AGHA cohort if they met at least one of two criteria: 1) having one or more first or second-degree relatives with documented cardiomyopathy or sudden death before the age of 50; or 2) if the patient is diagnosed before 60 years of age and has conduction disease or out of hospital cardiac arrest. Accessible phenotypic data included patient characteristics including sex, self-reported ethnicity, height, weight, age of diagnosis, symptoms, comorbid conditions, family history of cardiomyopathy and sudden death before 50 years of age, and several cardiac MRI measures and Holter monitor results. The data also included results from clinical genetic testing conducted by the AGHA cardiovascular flagship team to identify P/LP variants in known cardiac disease genes, as previously described [32]. Briefly, both Tier 1 genes (which have definite clinical validity) and Tier 2 genes (lesser-evidenced research-based genes) were analysed and identified variants were annotated based on the American College of Medical Genetics (ACMG) guidelines [33]. Genetic test results provided to us by AGHA were annotated into 3 categories: 1) Genotype +ve (G+): where a P/LP variant was identified; 2) VUS: where variant(s) of unknown significance were detected which may be associated with the patient’s condition but lack strong evidence for pathogenicity, and 3) Genotype -ve (G-): where no variant had been identified. VUS were further classified into three sub-groups: a) high clinical significance but not enough information to classify them as likely pathogenic, b) insufficient evidence to classify it as either disease causing or likely benign, and c) low clinical significance, the evidence suggesting they are likely to be benign. Where a P/LP or VUS variant had been identified, the location of the variant was provided as part of the participant phenotype data.

**Table 1:** Description of DCM patients from AGHA cohort.

|  |  |  |
| --- | --- | --- |
| **Proportion males** | 55% |  |
| **Mean diagnosis age in years (SD)** | 38.3 (15.8) |  |
| Males | 36.93 years (15.46) |  |
| Females | 40.07 years (16.18) |  |
| **Genetically determined ancestry N (%)** |  |  |
| African | 2 (1.56%) |  |
| American | 1 (0.78%) |  |
| East Asian | 8 (6.20%) |  |
| European | 113 (87.59%) |  |
| South Asian | 2 (1.56%) |  |
| Excluded for missingness | 3 (2.32%) |  |
| **Genetic results from screening of Tier 1 and Tier 2 DCM genes** | **European-ancestry individuals** | **Non-European ancestry individuals** |
| **P/LP in *TTN* gene (*TTN*_G+)** |  |  |
| N (%) | 20 (17.7) | 3 (18.8) |
| Mean diagnosis age in years (SD) | 44.6 (9.0) | 21.6 (2.7) |
| Percentage with family history of disease | 90% | 100% |
| **P/LP in non-*TTN* gene (non*TTN*_G+)** |  |  |
| N (%) | 18 (15.9) | 2 (12.5) |
| Mean diagnosis age in years (SD) | 32.0 (15.3) | 35.1 (22.4) |
| Percentage with family history of disease | 72.2% | 100% |
| **VUS** |  |  |
| N (%) | 17 (15.0) | 3 (18.6) |
| Mean diagnosis age in years (SD) | 42.53 (15.6) | 26.75 (22.4) |
| Percentage with family history of disease | 70.6% | 66.7% |
| **No P/LP/VUS variant identified (G-)** |  |  |
| N (%) | 58 (51.3) | 8 (50.0) |
| Mean diagnosis age in years (SD) | 38.76 (17.6) | 36.42 (9.9) |
| Percentage with family history of disease | 55.2% | 50% |

### 2.3. AGHA genotype data processing and quality control

Raw whole genome sequence data for 429 AGHA individuals were obtained through Australian Genomics and analysed through a variant calling pipeline based on the Genome Analysis Tool Kit (GATK) best practices [34]. Briefly, after initial quality check by FastQC (v0.11.9) [35] and quality control by Fastp (v0.23.2) [36], all sequencing reads were aligned to hg37 (GRChr37) using BWA-MEM algorithm within the Burrows-Wheeler Aligner (BWA) package (v0.7.17) [37]. The duplicate reads were identified and removed by Picard (v2.25.1). Single nucleotide variants (SNVs) and small indels were identified using GATK (v4.3.0.0) Haplotype Caller tool [38]. This was done by calculating metrices based on 13 known variant location including dbSNP, HapMap, 1000 Genomes, hg19 reference genome etc. After variant calling for the AGHA cohort, the genetically inferred sex of each individual was checked with self-reported gender and no mismatch found. The genetic ancestry of each individual was estimated by comparing the first two principal components (PCs) with those of the participants from the 1000 genomes study [39], with individuals classified into five groups: American (AMR), African (AFR), East Asian (EAS), South Asian (SAS) and European (EUR). As a basic genotype quality control step, samples with more than 1% missing data and SNPs with more than 1% missingness have been removed. This excluded 10 samples and 344,342 SNPs. The final dataset comprised 126 DCM patients (89.6% European ancestry). As the DCM GWAS is based on European-ancestry individuals only, all PRS analysis were restricted to European-ancestry individuals (113 DCM patients).

### 2.4. Reference population – The UK Biobank cohort

The UKB cohort was used as a population reference for polygenic risk score analysis. It consists of over half a million participants from the UK, having age between 40 to 69 years and recruited between 2006 to 2010 [40]. Genomic data along with deep phenotypic data were collected from participants at different time points. Consent was collected from all participants. The current study was conducted using the UKB Resource under Application Number 12505. This research is covered by The University of Queensland Human Research Ethics Committee approval (HREC number 2020/HE002938). The UKB participants who have withdrawn from the study as of 17 December 2024 were excluded at first and only the individuals of genetically determined European ancestry as described by Yengo et al [41] were included in this analysis. SNPs with more than 1% missingness have been removed. Thus, we included 450,280 UKB samples in this study. For evaluating the performance of different PRS methods, DCM cases were identified in UKB individuals based on death registry, self-reported illness and I42.0 billing code from the International Classification of Diseases 10^th^ edition (ICD-10) [4]. Using this definition, 1,503 DCM cases were identified in the UKB samples.

### 2.5. DCM GWAS Summary statistics

The DCM GWAS summary statistics were obtained from Zheng, S.L. et al. [14]. In the main study, a case-control meta-analysis was performed on 14,256 DCM cases and 1,199,156 controls from 16 studies of European participants, including UKB participants. In our study, UKB data is used as a reference sample for PRS analysis. Summary data from a meta-analysis of 7,743 DCM cases and 750,286 controls that excluded UKB participants were provided by the authors through the HERMES consortium for generating a DCM PRS.

### 2.6. Generating polygenic risk scores

PRS is the combined effect of the disease risk variants that an individual carries, reflecting the genetic predisposition of an individual for a particular disease. PRS is calculated as the weighted sum of disease risk allele counts, whereby for *n* disease-associated variants, 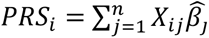, where *X_ij_* represents the genotype for the *i^th^* individual coded as 0, 1 or 2 depending on the allele dosage for *j^th^* SNP. 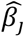 is the magnitude of the per-allele effect on disease risk for *j^th^* SNP. Different methods for PRS calculation use different approaches for selecting the disease-associated variants and estimating 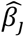 from GWAS summary statistics. Simpler approaches to PRS calculation only consider independently associated genome-wide significant SNPs (with *p* < 5 × 10^-8^) and effects estimated in the original GWAS. However, the ability to identify variants at this significance threshold is dependent on sample size, and the inclusion of a larger number of SNPs with weaker association p-values has been shown to lead to a more powerful PRS for polygenic traits [42], while more sophisticated methods incorporate variant functional annotation to re-estimate effect sizes [43]. There are few other issues to be considered to increase the PRS accuracy that include consideration of the ancestry of the target participants, incorporation of effect sizes from different ancestral background, accounting for the mixture of multiple distribution of varying effect size, inclusion of functional genomic annotation etc. Bayesian approaches have shown to be more powerful, and they do not require additional training dataset for SNP selection and weight estimation [44]. In this study, we generated PRS using commonly-used Bayesian methods: SBayesRC [43], SBayesR [44], LDpred2-auto [45], LDpred-funct [46] and PRScsx [47]. SBayesR extends the ridge regression by modelling effect sizes with a mixture of normal distributions. SBayesRC is an extension of SBayesR and incorporates functional genomic annotation for more than 7 million high-density SNPs. This method implements a low-rank approximation using the eigenvalues and eigenvectors of block-wise linkage disequilibrium (LD) correlation matrices. LDpred2-auto adjusts the effect size by accounting for the LD and automatically infers the fraction of causal variants. LDpred-funct is an extension of LDpred2-auto that incorporates functional annotations to prioritize the causal variants and refine the effect sizes. PRScsx integrates population specific architectures and cross-ancestry information, aiming to improve the predictive performance of the score. The variants and corresponding weights from each method were used to generate PRSs for each individual using PLINK (v1.9) [48]. The methods regarding each PRS are mentioned in the supplementary methods.

### 2.7. Evaluating polygenic risk scores in UK Biobank

To identify the best performing PRS method, we grouped the UKB participants into two equal sets, stratified by their sex and DCM status. Thus, we obtained training and testing sets of 225,140 participants each. The association of the PRS generated from each of the five abovementioned methods with DCM diagnosis was evaluated in the UKB test set using logistic regression, with age at enrolment, sex and the first 10 genetic principal components (PC) as covariates. The odds ratio for DCM per standard deviation (SD) increase in PRS is reported along with the proportion of variance in disease explained by the PRS using the Nagelkerke’s pseudo-R^2^ (calculated using the rcompanion R package). For this, the Nagelkerke’s pseudo R^2^ was first estimated for the base model, with only the covariates as predictors, followed by a full model with PRS and covariates. The difference in the Nagelkerke’s pseudo R^2^ between the base and full model is the proportion of variance explained by the PRS alone.

### 2.8. Comparing PRS in the AGHA DCM cohort with the UK Biobank population sample

Given previous evidence of disease risk conferred by a PRS in the top 10% of the population distribution being similar to monogenic variants for several cardiovascular diseases, AGHA individuals with a PRS within the top 10% of the UKB reference population sample DCM PRS distribution were considered to have a high polygenic burden. We used the full UKB set as the reference population sample (450,280 European samples). Using the PRS method which showed the best prediction accuracy in UKB test set, the DCM PRS were calculated for the AGHA DCM cases. The PRS distributions in the G+ and G- AGHA individuals were compared to the UKB reference population distribution using a Mann-Whitney U test. Given the higher prevalence but lower penetrance of *TTN* variants, G+ individuals were further stratified by the presence of a P/LP variant in the *TTN* gene (*TTN*_G+ and non*TTN*_G+). One individual who had a P/LP variant in the *TTN* gene also had a P/LP variant identified in the *MYH7* gene and was grouped within the non*TTN*_G+ group. AGHA individuals with a PRS in the bottom decile of the UKB reference PRS distribution were also identified. The Fisher’s exact test was used to test whether the proportion of AGHA DCM patients in each genotype group that fell in the top and bottom PRS deciles were significantly different than those observed in the reference population, where the expected proportions are 0.1 and 0.9 for being the top decile versus not, or for being in the bottom decile versus not.

### 2.9. Integrating multiple PRS methods using PRSmix

The performance of the polygenic risk scores in identifying individuals with high disease burden depends on the underlying method, the baseline GWAS, phenotype definition and the training dataset. Although different methods applied to the same GWAS data generate highly correlated PRS, classification of the individuals into different risk groups, especially towards the extreme ends of population distribution, tends to be unstable [49]. The PRSmix method [50] combines different PRS into an integrated risk score using an elastic net regression model on training data to produce a weighted linear combination of all input PRS. It has been reported that this integrated score provides stable and consistent classification for high-risk individuals [51]. The UKB training set was used to estimate the weights for each PRS method along with their age, sex and first 10 PC of genetic ancestry as covariates. All other default parameters for PRSmix were used. PRSmix score was generated for UKB and AGHA participants using these estimated scores.

### 2.10. Agreement between SBayesRC and PRSmix

To evaluate the consistency between SBayesRC and PRSmix, the PRS of the UKB test samples was divided into 100 equal bins, and each study sample was classified into a percentile bin based on the PRS range of each bin. We used Jaccard Index [52], which is the measure of similarity among two sets, to evaluate agreement in placing a particular individual into the top or bottom decile of the UKB testing sample PRS distribution.

## 3. Results

### 3.1. Evaluation of DCM PRS methods in UK Biobank

All PRS were significantly associated with increased risk of DCM in UKB, with SBayesRC showing the strongest association (Odds ratio per 1 SD increase in PRS = 1.72 [95% CI, 1.58 – 1.88] and Nagelkerke’s R^2^ of 0.017). As the best performing individual method, SBayesRC was used in subsequent analysis.

### 3.2. Comparison of PRS in AGHA DCM patients and the UK Biobank population reference sample

As expected, compared to the UKB population reference samples, the PRS in the AGHA G-DCM patients was statistically significantly higher (median PRS 0.47 vs 0.32 and Bonferroni adjusted p = 0.03). Interestingly, the *TTN*_G+ AGHA DCM patients also had a statistically significantly higher PRS than UKB individuals (median PRS 0.64 vs 0.32; Bonferroni adjusted p = 0.0006) and had the highest median PRS out of all groups, while the PRS in the non*TTN*_G+ AGHA DCM patients was not statistically different to UKB (median PRS 0.36 vs 0.32; Bonferroni adjusted p = 1.0) (Figure 3).

**Figure 2.**
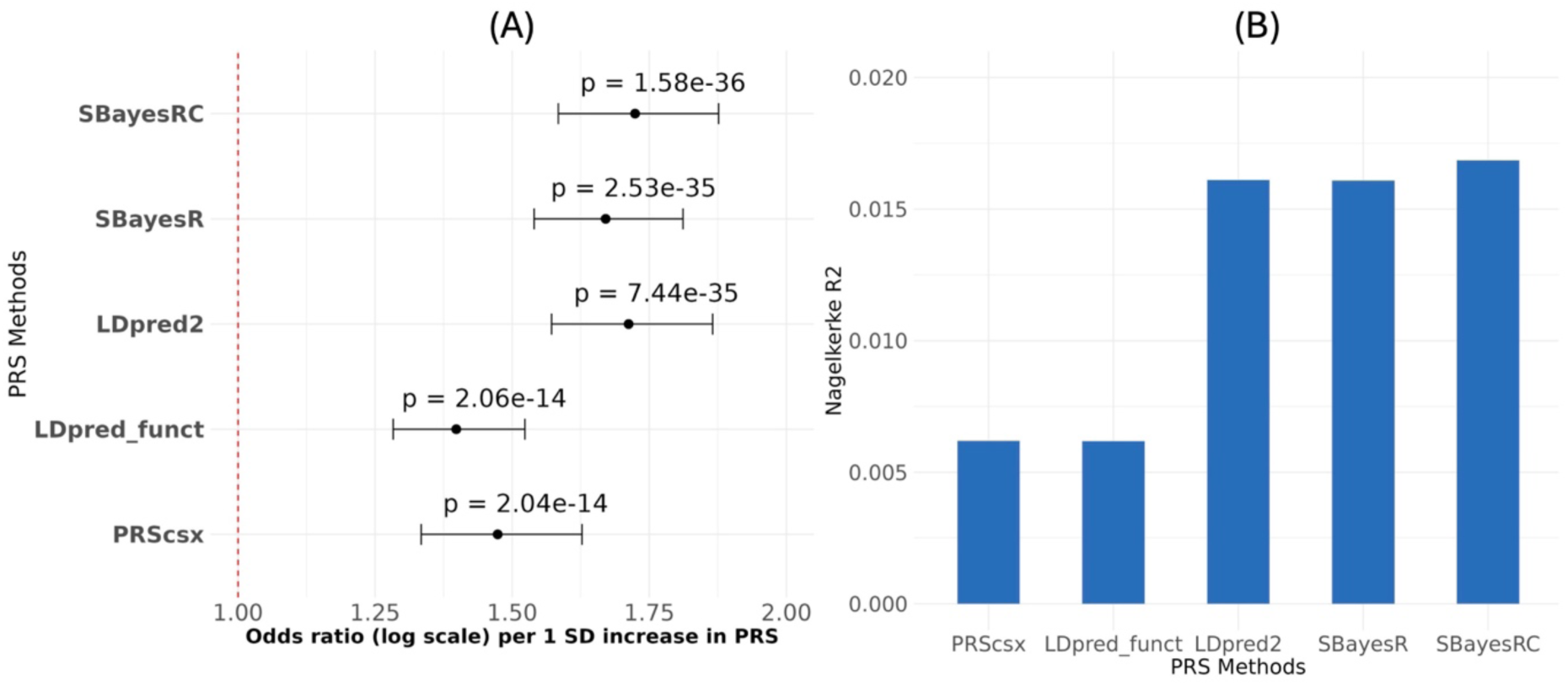
Comparison of different PRS methods for predicting DCM using (A) odds ratio (OR) for DCM per SD increase in PRS and (B) Nagelkerke’s R^2^.

**Figure 3:**
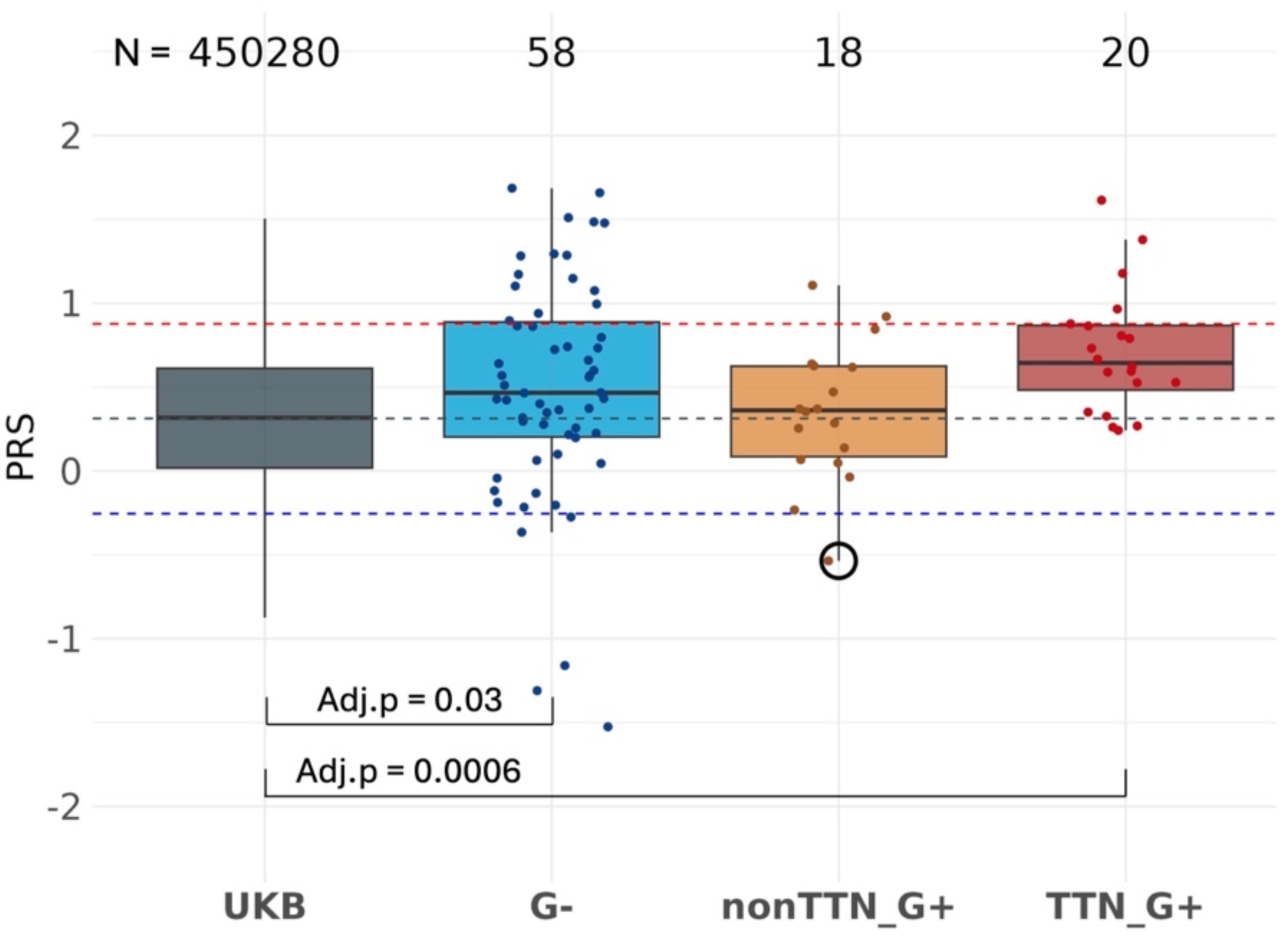
The mean DCM PRS of both G-and *TTN*_G+ groups were statistically significantly higher than the mean DCM PRS of UKB participants. There was no significant difference between mean DCM PRS of other groups. The grey horizontal dashed line indicates the mean PRS value, and the red and blue lines represent the PRS values corresponding to the top and bottom deciles of the UKB population reference distribution, respectively.

### 3.3. An enrichment of AGHA DCM G- individuals in the top decile of the reference population PRS distribution

The proportion of individuals with a PRS in the top decile was significantly higher in G-individuals (26%, N = 15, p = 0.0005) and *TTN*_G+ individuals (25%, N = 5, p = 0.04) relative to the reference population (Figure 4). No enrichment in individuals with a PRS in the top decile was observed in the non*TTN*_G+ group (11%, N = 2, p-value = 0.69). While nine percent (N = 5) of the G- individuals and 6% (N = 1) of the non*TTN*_G+ individuals had a PRS in the bottom decile, none of *TTN*_G+ individuals had a PRS in the bottom 4 deciles of the population reference distribution (Figure 4).

**Figure 4.**
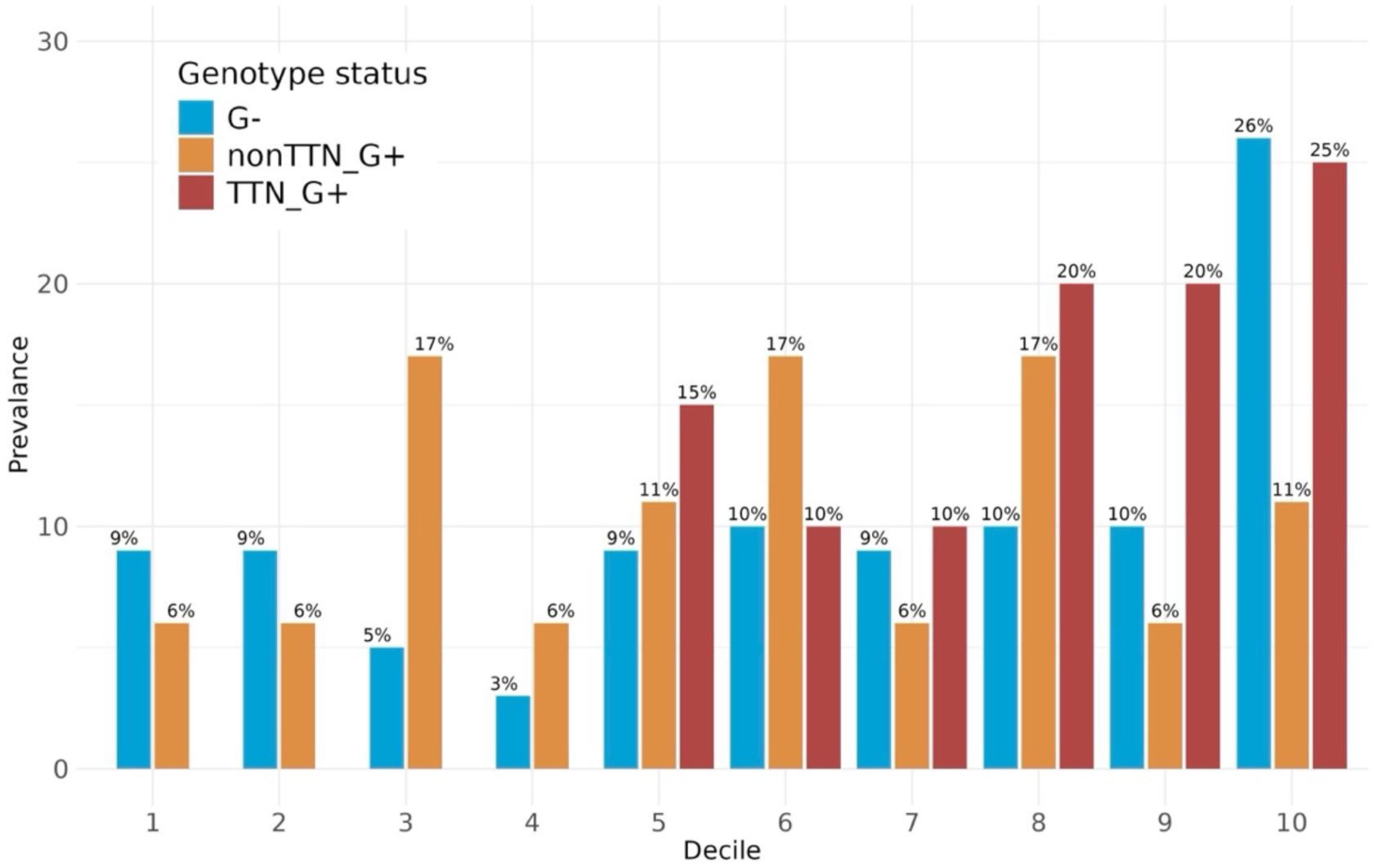
Number of AGHA samples in different genotype groups, having PRS that falls in different deciles of UKB distribution identified by different PRS methods. Percentage of G-, *TTN*_G+ vs non*TTN*_G+ patients identified in different risk groups.

### 3.4. High polygenic burden may contribute to disease development in *TTN* variant carriers

One potential explanation for the higher PRS burden in *TTN*_G+ patients may be the lower penetrance of *TTN* variants and therefore the requirement of additional polygenic burden to cross the disease laibility threshold. Alternatively, variants included in the PRS could be in LD with the *TTN* P/LP variants and thus lead to an inflation in the PRS. To test the latter hypothesis, the DCM PRS was recalculated excluding any variants within 10Mb upstream and 10Mb downstream of the *TTN* gene. No significant difference was observed between the original PRS and the PRS generated by excluding all variants around the *TTN* gene (Figure 5). These results support the hypothesis that additional genetic risk burden may lead to disease development in *TTN* variant carriers. Interestingly, the individual with both a P/LP variant in the *TTN* and *MYH7* genes had a PRS in the bottom decile. Therefore, additional genetic risk burden amongst affected *TTN* variant carriers could be attributed to both high polygenic burden or additional monogenic variants. However though given the low prevalence of monogenic variants, a high polygenic burden would be expected to contribute to genetic risk in a greater proportion of affected *TTN* variant carriers.

**Figure 5:**
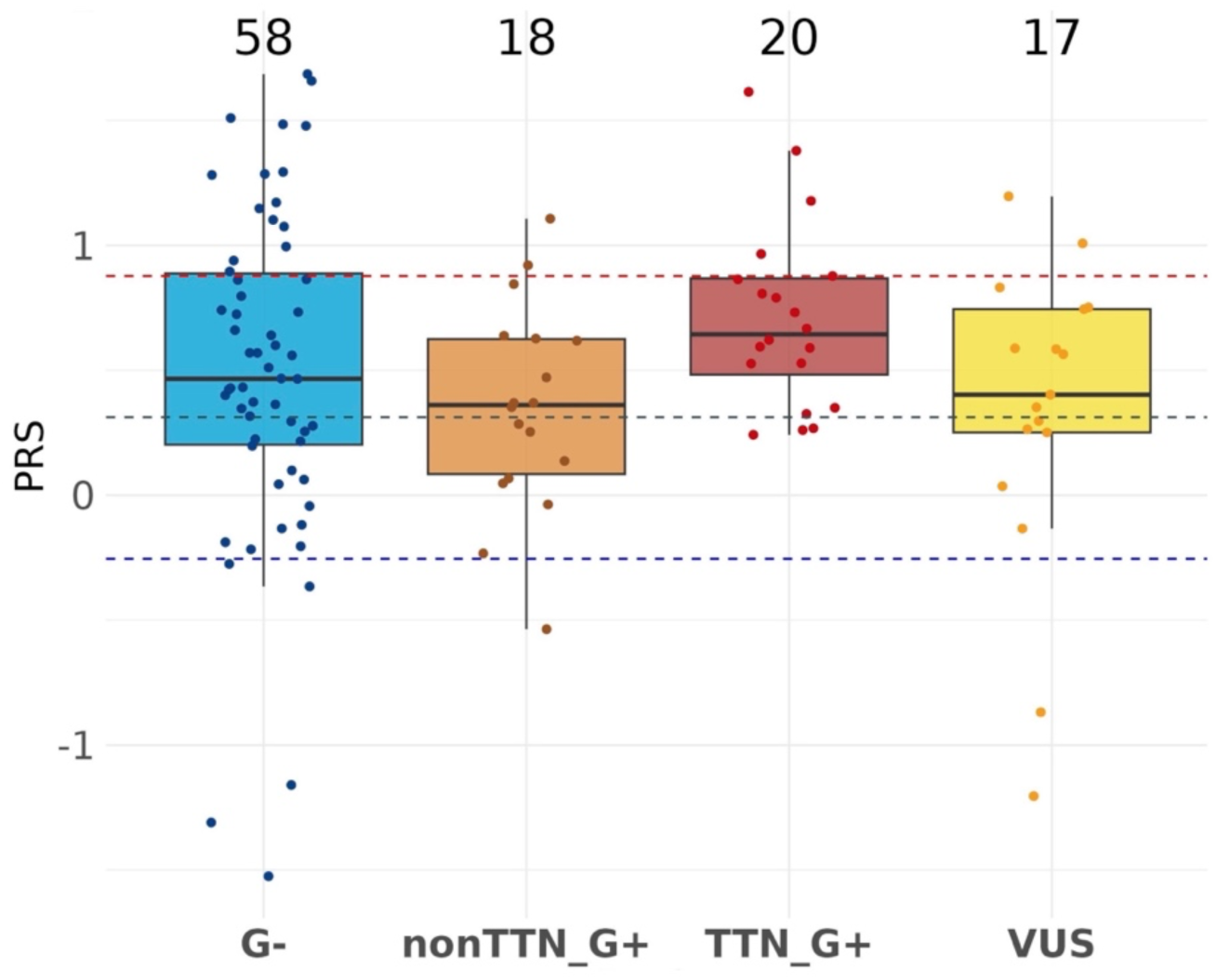
DCM PRS without variants in *TTN* gene region among different pathogenic groups of AGHA DCM patients. There is no statistically significant difference among the groups. The grey horizontal dashed line indicates the mean PRS value, and the red and blue lines represent the PRS values corresponding to the top and bottom deciles of the UKB population reference distribution, respectively.

### 3.5. Further investigation of individuals with very low PRS

Three G- individuals and two individuals with a VUS had PRS within the bottom decile (Figure S1). All had a family history of disease, which increases the likelihood of monogenic disease variants. Amongst the three G- individuals, one had an age-of-onset <5 years. Amongst the individuals with a VUS, one carries a VUS in the *TTN* gene and another carries a VUS in the *LMNA* gene. The very low PRS warrants more comprehensive whole-genome sequence analyses amongst the G- to identify novel monogenic variants, while functional studies on the *LMNA* VUS may help with reclassification of this variant to P/LP. As mentioned previously, one individual with a *TTN* P/LP variant had a PRS in the bottom 5% of the reference population distribution but carries an additional P/LP variant in the *MYH7* gene. Given this observation, more comprehensive genetic analysis for the identification of a second, novel monogenic variant may be warranted in the individual with a *TTN* VUS with a PRS in the bottom decile.

### 3.6. Consistency in PRS methods

A potential application of PRS in DCM genetic testing is for identifying G- individuals with very high PRS who therefore have a likely polygenic cause for their disease or identifying G-individuals with a very low PRS for more comprehensive genomic analysis for novel variant identification. Therefore, confidence in identifying individuals with very low or high PRS is crucial. Consistency among different PRS methods for ranking individuals has been a concern for clinical utility [53]. PRSmix [50] is a method that was developed to overcome this issue by combining multiple PRS methods to improve risk stratification. Overall, there was a strong correlation in assigning individuals to PRS percentiles between SBayesRC and PRSmix (Figure 6), especially at the highest and lowest percentiles. The Jaccard index between SBayesRC and PRSmix for selecting individuals in top decile was 0.83 (i.e. 83% overlap between the two methods) for both *TTN*_G+ group and G- group, and 0.67 for the non*TTN*_G+ group. This increased to 0.91 in the G- group when considering individuals in the top 20% of the PRS distribution. In the case of selecting individuals in bottom decile, the Jaccard index between SBayesRC and PRSmix was 0.6 for G- group.

**Figure 6:**
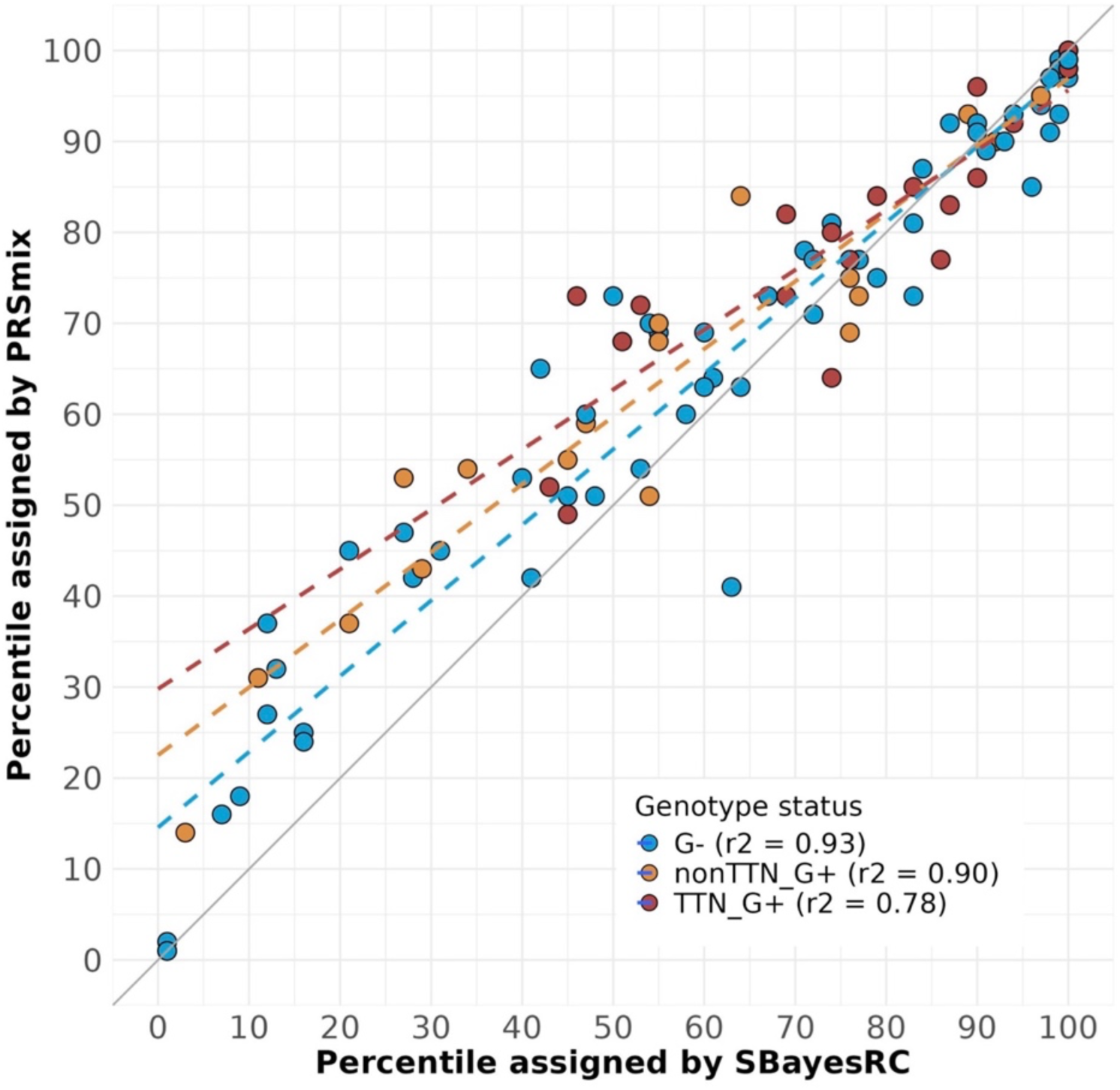
Concordance among SBayesRC and PRSmix in categorizing DCM cases in different PRS percentile of population reference distribution. The grey solid line is the identity line, and the dashed lines indicate the regression line for corresponding colour groups.

## 4. Discussion

Despite recent advances in our understanding of the polygenic contribution in DCM, studies evaluating the application of PRS as part of genetic testing for DCM are lacking. This study assessed PRS amongst DCM patients who had undergone clinical genetic testing for monogenic variant identification to determine whether PRS could inform clinical management of affected patients and at-risk family members.

The results demonstrate that integration of PRS analysis has the potential to increase the diagnostic yield of genetic testing, as well as provide a more comprehensive view of the genetic contribution to disease risk, which in turn can inform risk stratification. The proportion of G-DCM patients with a likely polygenic cause of disease has not been quantified before. Here we show that 26% of G- DCM patients had a PRS in the top decile. In these individuals a high PRS may be the primary genetic contributor to disease susceptibility. However, further data are necessary to understand the likelihood of novel monogenic variants in G- DCM patients with a high PRS before the latter can be confidently used for diagnostic genetic testing in clinical settings.

An unexpected finding from this study was the notable difference in the PRS distribution in *TTN* and non-*TTN* P/LP variant carriers. Whilst the PRS in non-*TTN* P/LP carriers was not found to be significantly different from the UKB population reference distribution, DCM individuals with *TTN* P/LP variants had the highest mean PRS (which was statistically significantly higher than UKB), with 25% of *TTN* P/LP variant carriers amongst the top decile of the reference PRS distribution, as well as a complete depletion of *TTN* variant carriers in the lowest four PRS deciles. The sample sizes in these two groups are small, and further validation in larger cohorts is necessary to confirm this difference in polygenic burden in the *TTN*_G+ and non*TTN*_G+ groups. However, given the *TTN*_G+ and non*TTN*_G+ groups had similar samples sizes, the depletion of low PRS in the *TTN*_G+ group is striking. This could potentially explain variable expressivity among family members carrying the same *TTN* variant. Thus, in addition to cascade testing in at-risk relatives of affected *TTN* variant carriers, knowledge of PRS could potentially inform risk stratification in family members and inform tailored clinical screening strategies.

The most recent European Society of Cardiology guidelines on cardiomyopathy screening do not recommend cascade genetic testing in family members if a P/LP has not been identified in an affected individual [12]. Among AGHA participants, three G- and two VUS DCM patients had a PRS in the bottom decile of the population distribution. Based on previous studies that have demonstrated an enrichment of pathogenic variants in affected individuals with very low PRS [31], as well as considering patient characteristics of these 5 individuals, further genetic investigation would be warranted for identification of novel monogenic variants before ruling out cascade genetic testing in family members. Additionally, recent studies have demonstrated that polygenic scores are largely similar between siblings and therefore high PRS can indeed segregate within families [54]. Using UKB data, Reid et al. showed that in individuals with a high PRS for CAD (defined as the top 20% of the distribution), 44% of siblings’ polygenic scores also fell within the top 20% [54]. This positive predictive value of a high polygenic score was largely independent of family history of disease. Similarly, PRS analysis for 24 common diseases in the FinnGen study showed that the average concordances of a high PRS among first-degree relatives was 33.7% [55], and that PRS explained on average 10% of the effect of first-degree family history. Therefore, cascade testing in first-degree relatives, in particular siblings of G- individuals with a high DCM PRS may warrant consideration.

Given the small sample size of the AGHA cohort, validation of these observations in larger DCM cohorts and also in DCM cohorts enriched for sporadic DCM cases is necessary. All analyses were restricted to European ancestry individuals due to the lack of non-European individuals in the DCM GWAS participants and the resulting lack of a well-powered DCM PRS for non-European individuals. This is a major caveat in almost all genetic studies that can only be addressed by increasing the participation of individuals from diverse ancestries in genetic research. With several ongoing efforts such as All of US [56], China Kadoorie Biobank [57] and Million Veteran Program [58] to diversify genomic data, such analyses will likely become feasible in other populations in the near future. Moreover, it is well established that the UKB exhibits a healthy-volunteer bias, with people who have common cardiovascular conditions being underrepresented compared to the general population [59], suggesting that the results should be interpreted with caution. While our study demonstrates strong concordance between two of the best performing PRS methods for identifying individuals with very high or low PRS, future studies should also explore different PRS methods.

As PRS methodology matures and larger DCM GWAS become available in the near future, PRS accuracy and confidence in their clinical application will increase. Currently, PRS is limited to estimating an individual’s genetic liability to disease based on common variants. As the same P/LP variants may only be observed in very few individuals, it is not currently possible to estimate their magnitude of effect on disease risk, and thus such variants cannot be incorporated into PRS calculations. However, as more whole genome sequence data become available, development of PRS that incorporates the full frequency spectrum of risk variants may become feasible. A complete picture of an individual’s genetic risk, in combination with non-genetic risk factors, could facilitate more tailored clinical management of affected individuals and their family members.

## Data Availability

All data used are publicly available:
https://www.ukbiobank.ac.uk/use-our-data/apply-for-access/
https://www.australiangenomics.org.au/tools-and-resources/accessing-australian-genomics-data/

## Supplementary methods

### Polygenic risk score generation

We used five different methods for generating polygenic risk score (PRS) that includes SBayesR, SBayesRC, LDpred2 auto, LDpred_funct, PRScsx and PRSmix.

#### 1.1. SBayesR

A linkage disequilibrium (LD) reference dataset was downloaded from the SBayesR website (https://gctbhub.cloud.edu.au/software/gctb/#SBayesRTutorial) (banded LD matrix) and default settings were used unless otherwise stated (--exclude-mhc; -- chain-length 10000; --burn-in 2000; --no-mcmc-bin).

#### 1.2. SBayesRC

SBayesRC is an extension of SBayesR that incorporates the functional genomic annotations with high density SNPs (>7 millions). The functional annotations were downloaded from the SBayesRC website (https://gctbhub.cloud.edu.au/software/gctb/#SBayesRCTutorial). The eigen-decomposition data European ancestry was also downloaded from the website. Before using executing the SBayesRC, the summary statistic was imputed for the 7 million SNPs using GCTB software.

#### 1.3. LDpred2 auto

For LDpred2, we estimated the SNP weights using the automatic model (LDpred2 auto) as it does not require any validation set to infer the hyperparameters. We used the bigsnpr R package for this. We restricted our analysis to the HapMap3 SNPs and downloaded the LD reference from the vignettes of the R package (https://privefl.github.io/bigsnpr/articles/LDpred2.html).

#### 1.4. LDpred funct

We used the python package cloned from the github page (https://github.com/carlaml/LDpred-funct). At first, we estimated the per-SNP heritability using the ldsc software (https://github.com/bulik/ldsc) under the baseline LD. The baseline LD annotations were downloaded from the github page of LDpred funct. All the analyses were restricted to HapMap3 SNPs.

#### 1.5. PRScsx

PRScsx integrates GWAS summary statistics and external LD reference panels for multiple ancestry population. But we used only European population, which makes this similar to PRScs. We used the LD reference panels constructed using the UK Biobank data for European samples downloaded from the github page of the software (https://github.com/getian107/PRScsx).

## Supplementary figures

**Figure S1:**
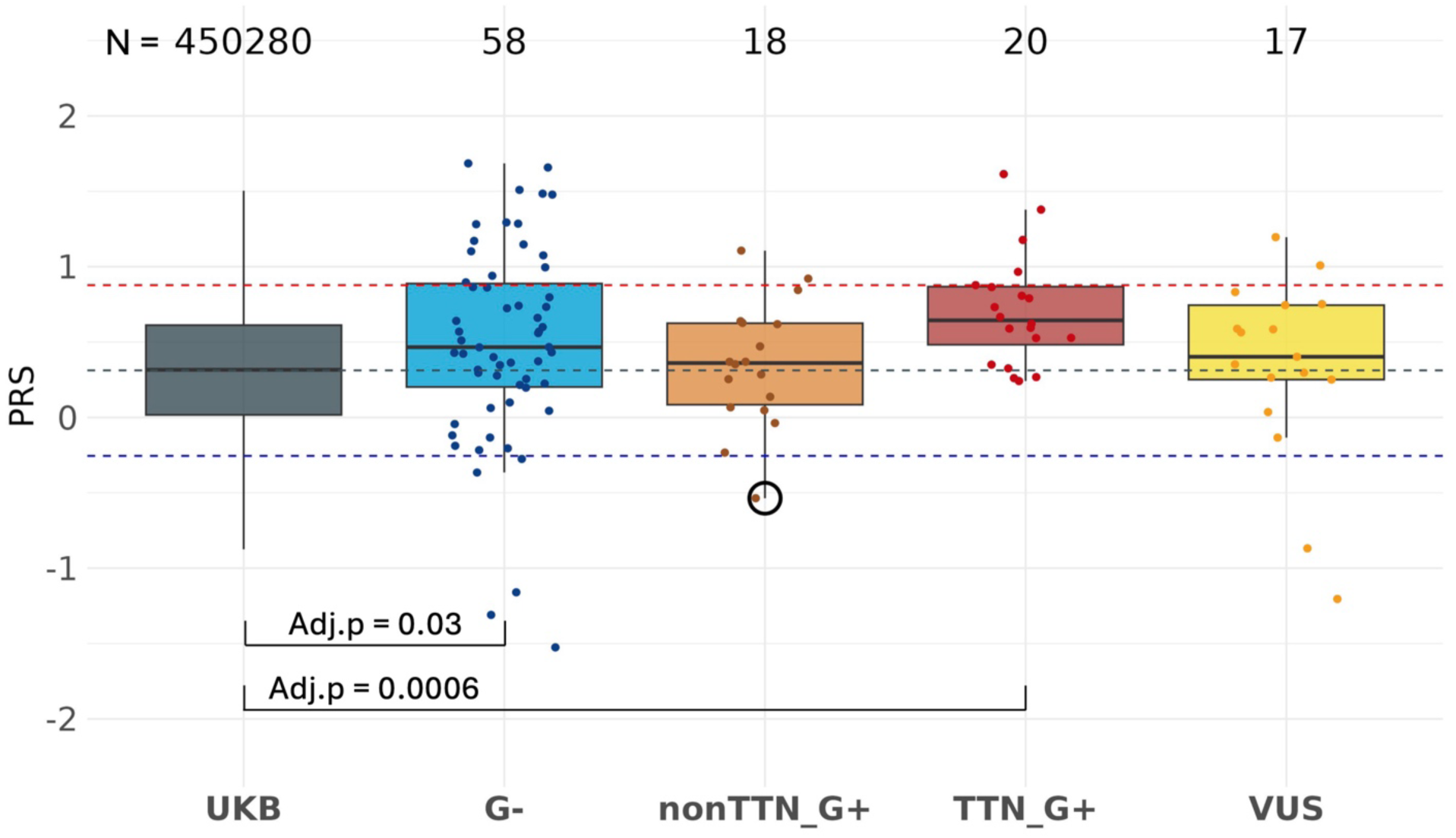
PRS distribution among different pathogenic groups of the AGHA participants, compared to UKB participants. Three samples with G- and 2 individuals with VUS have very low PRS.

## Supplementary tables

**Table S1:** Description of the data for the AGHA samples. DCM – Dilated cardiomyopathy, HCM – Hypertrophic cardiomyopathy, RCM – Restrictive cardiomyopathy, LVNC - Left Ventricular Non-Compaction, ARVC – Arrhythmogenic right ventricular cardiomyopathy, LQTS – Long-QT syndrome, CPVT - Catecholaminergic polymorphic ventricular tachycardia, and BrS – Brugada syndrome.

| **Description** | **Total N – Mean**<br>**diagnosis age in**<br>**years (SD)** | **Female N (%) – Mean**<br>**diagnosis age in years**<br>**(SD)** | **Male N (%) – Mean**<br>**diagnosis age in years**<br>**(SD)** |
| --- | --- | --- | --- |
|  | 429 – 35.63 (16.10) | 195 (45.45%) – 36.72<br>(16.0) | 234 (54.55%) – 34.72<br>(16.17) |
| **Cardiomyopathies** |  |  |  |
| DCM | 129 – 38.34 (15.80) | 58 (44.96%) – 40.07<br>(16.18) | 71 (55.04%) – 36.93<br>(15.46) |
| HCM | 143 – 34.82 (14.92) | 40 (27.97%) – 34.42<br>(14.49) | 103 (72.03%) – 34.98<br>(15.15) |
| RCM | 3 – 7.97 (6.27) | 2 (66.67%) – 11.33 (3.3) | 1 (33.33%) – 1.25 (-) |
| LVNC | 21 – 27.79 (19.68) | 7 (33.33%) – 30.18<br>(20.7) | 14 (66.67%) – 26.60<br>(19.84) |
| ARVC | 33 – 39.83 (12.47) | 14 (42.42%) – 41.19<br>(14.19) | 19 (57.58%) – 38.83<br>(11.35) |
| **Arrhythmia Syndromes** |  |  |  |
| LQTS | 81 – 33.91 (17.03) | 66 (81.48%) – 36.70<br>(15.91) | 13 (16.05%) – 22.94<br>(17.86) |
| CPVT | 9 – 26.80 (18.03) | 2 (22.22%) – 30 (28.28) | 7 (77.78%) – 25.88<br>(17.72) |
| BrS | 10 – 44.97 (13.18) | 4 (40%) – 35.23 (9.55) | 6 (60%) – 51.46<br>(13.18) |

